# Corticosteroids reduce vascular ultrasound sensitivity even with short duration: results from Coventry Multi-Disciplinary Fast Track Pathway for management of Giant Cell Arteritis

**DOI:** 10.1101/2020.10.17.20214213

**Authors:** Jonathan Pinnell, Purnima Mehta, Carl Tiivas, Shirish Dubey

## Abstract

**Introduction:** Giant cell arteritis (GCA) is a common form of vasculitis and can result in permanent visual loss and other complications. The advent of vascular Doppler ultrasound (US) has provided a new means for early diagnosis for these patients although it is affected by introduction of corticosteroids (CS). The Coventry multidisciplinary fast track (FTGCA) pathway was set up in 2013 in collaboration with vascular physiology and ophthalmology with a view to enabling prompt multidisciplinary assessment.

**Objectives:** This study aims to assess feasibility of this novel pathway and to assess the impact of CS use on the performance of US in a real life cohort.

**Methods:** Data were collected retrospectively for patients who attended the Coventry FTGCA pathway between 1st Jan 2014 to 31st December 2017. Patients were identified from US lists and clinical details were obtained from electronic medical records. Ethical approval was obtained from Research and Development department.

**Results:** 620 eligible patients were included in this study. The pathway overall performed well with significant reduction in patients needing CS. US had sensitivity of 50% which improved further to ∼56% in CS naïve patients although median duration of CS use was 2 days. US specificity was >96%, and we were able to avoid using CS completely in 451 patients (73%). CS negatively impacted on utility of US with US more likely to be false negative.

**Conclusions:** This novel pathway demonstrates the ability to minimise use of CS through fast track multidisciplinary assessment. US was performed promptly and had reassuring real life sensitivity and specificity in this cohort. CS naïve patients showed significantly higher sensitivity for US despite the short duration of CS use.

## Introduction

Giant cell arteritis (GCA) commonly presents with headache in the temporal region, scalp tenderness, jaw claudication and visual disturbance [1]. It is the most common type of vasculitis affecting 22 per 100,000 people aged over 50 in the UK and is characterised by systemic inflammation, arteritis and critical ischaemia [2]. It is often associated with polymyalgia rheumatica which causes pain and stiffness in the proximal muscle groups [1]. Permanent loss of vision, resulting from ischaemic optic neuropathy and retinal ischaemia is the most feared complication of GCA and can affect up to one fifth of patients [3]. Strokes and aortitis leading to aortic aneurysms are recognised later stage complications, but visual loss tends to occur early making GCA a medical emergency [4]. Permanent visual loss leads to long term economic, health and social consequences for the patients, their family and society in general [5]. Consequently, British Society for Rheumatology (BSR) guidelines emphasise the importance of prompt recognition and management of GCA [2].

### Clinical challenges in GCA

The diagnosis of GCA is challenging due to the lack of specific symptoms and the urgency of recognition. Temporal artery biopsy (TAB) has traditionally been the gold standard for investigation despite its limitations of cost, rare skillset and diagnostic delay. Patients are typically commenced on high dose corticosteroids (CS) whilst awaiting biopsy results. Whilst TAB has high specificity, it lacks sensitivity as skip lesions, which have been well described, may cause biopsy samples to be inadequate or contain unaffected segments of the temporal artery [6]. Therefore, negative TAB results do not always exclude the diagnosis, adding to diagnostic uncertainty.

The emerging use of ultrasound Doppler (US) in the diagnosis of GCA has helped overcome some of the challenges of relying on TAB [7]. It can be performed quickly, inexpensively, and non-invasively. Typical findings of hypoechoic haloes, stenoses, or occlusions on temporal artery ultrasonography have a diagnostic sensitivity and specificity of 78% and 79% respectively when compared to TAB and 87% and 96% when compared to ACR classification criteria for GCA [7-9]. Drawbacks of US include the need for skilled and experienced sonographers as well as concerns that the halo sign, the most diagnostic feature on US, can disappear within 48 to 72 hours of starting CS [10,11].

### Introduction of a multi-disciplinary fast track pathway

University Hospital Coventry and Warwickshire NHS Trust (UHCW) established a multi-disciplinary Fast Track Pathway (FTP) for GCA in 2013. It sought to address previously unmet needs by offering patients a same day clinical review, ultrasound Doppler, diagnosis, and treatment initiation for patients with GCA. This is the first GCA service to ask referrers to send the patient across for immediate assessment and investigations (during working hours) and without the need to start CS prior to assessment. Prior to the introduction of this service, patients experienced a delay of at least four weeks from initial presentation to diagnosis by TAB. The core aims of the FTP were: a) to avoid unnecessarily exposing unaffected patients to CS b) to minimise delays and anxiety for patients and c) to prevent avoidable GCA related vision loss by assessing and treating affected patients promptly. This pathway is the outcome of collaboration between the Rheumatology, Ophthalmology and Vascular Laboratory teams.

The aims of this study were:

1. Assess the feasibility of running a fast track pathway without prior introduction of CS
2. Assess the impact of the use of CS on performance of US in a real life cohort.

## Methods

### Description of the UHCW GCA FTP

Detailed description of the pathway is provided in another article [12]. Telephonic referrals to the fast track pathway are accepted if the patient is aged over 50 and has appropriate clinical features (see Figure 1). Patients are seen on the same day as the referral, or the next working day if they are referred out of hours. Those patients that are referred out of hours are usually commenced on CS immediately to avoid treatment delays. Comprehensive clinical assessments (including ophthalmic assessment), relevant blood investigations and vascular doppler ultrasound (US) studies are performed urgently and a management plan is instituted all on the same day. In patients with negative or inconclusive US and clinical suspicion, TAB is performed by the Ophthalmic team, usually within 2 weeks. Patients with low clinical suspicion of GCA and a negative US are reassured and discharged following the initial consultation. They undergo rapid CS tapering if they had been started. Patients with positive US are immediately commenced on oral CS or IV methylprednisolone if there is a risk of vision loss. IV methylprednisolone is delivered on the Rheumatology Day Unit where possible to avoid hospital admission, but rarely patients are admitted for inpatient treatment. In uncertain cases, CS are started/continued until more data are available.

**Figure 1:**
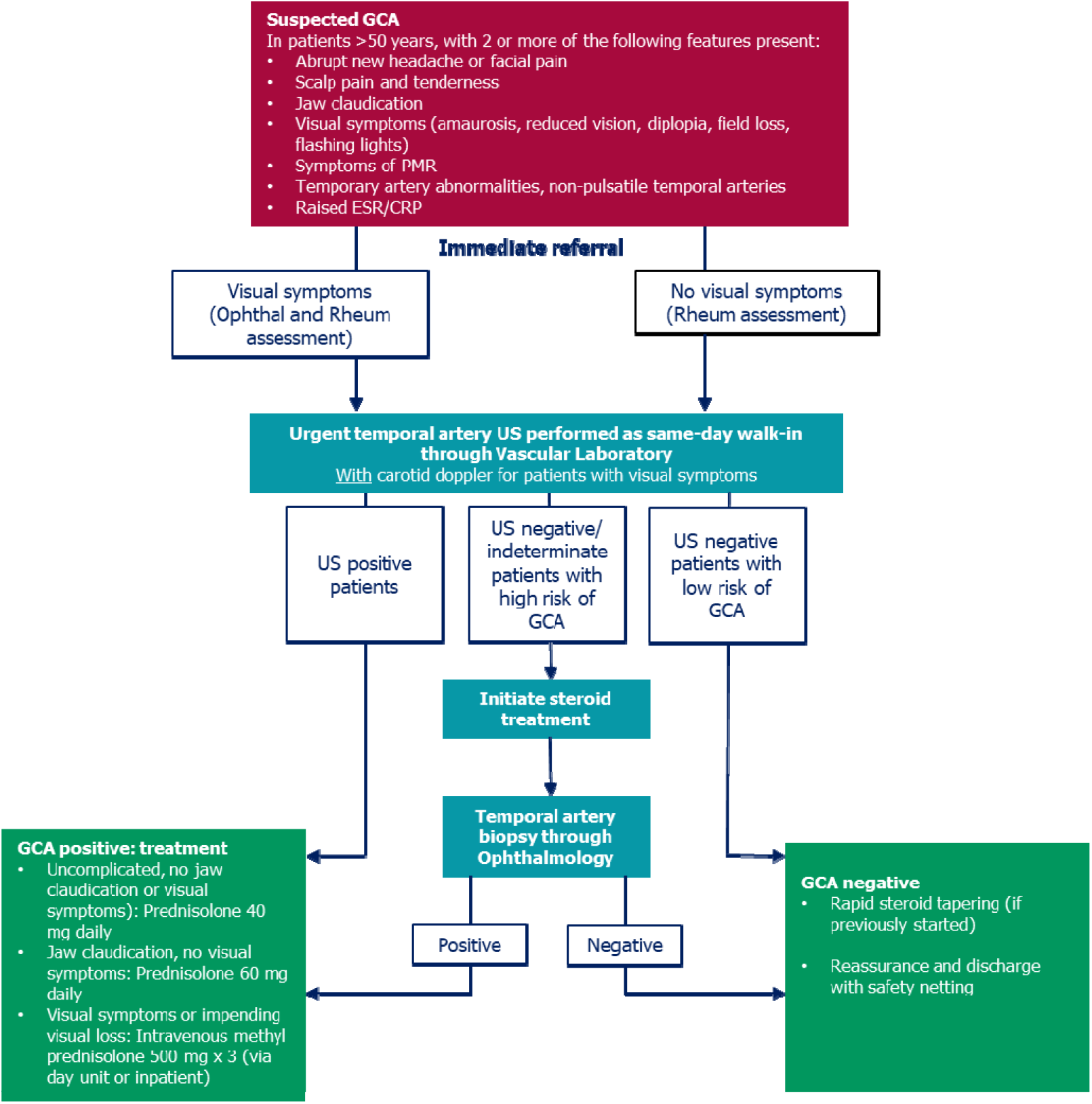
Coventry multi-disciplinary Fast-Track Pathway for Giant Cell Arteritis CRP: C-reactive protein; ESR: erythrocyte sedimentation rate; Ophthal: Ophthalmology; Rheum: Rheumatology; GCA: giant cell arteritis; PMR: polymyalgia rheumatica; US: ultrasound.

Our pathway started in middle of 2013 and initially we piloted this for a few months obtaining TABs alongside US with excellent results^12^. In this article, we present data from 1^st^ January 2014 to 31^st^ December 2017 – a period of 4 years. The data collected were collated to examine the impact on outcomes, reliability of US in the clinical setting, and the practical experience of running a fast track pathway.

Patients were identified from ultrasonography lists as all patients attending the fast track pathway are required to undergo an ultrasound Doppler of their temporal arteries. The clinical records of all patients with suspected GCA seen on the UHCW GCA Fast Track Pathway between, and including, January 2014 and December 2017 were retrospectively reviewed so that we could obtain data relating to final diagnosis and outcomes. A predetermined piloted proforma was used to collect data regarding patient demographics, clinical features, investigation results, CS use, and final diagnosis. All patients who had been referred with suspected GCA were included and missing data were sought from electronic and paper records. Patients who were less than 50 years old were excluded from this analysis.

Descriptive analyses were performed using Microsoft Excel and freely available online calculators were used for Fisher’s exact test to prove statistical difference. Sensitivity, specificity, positive predicted values and negative predicted values were estimated, together with 95% confidence intervals (CIs) using Wilson’s method [13]. Ethical approval was granted by the UHCW Research and Development Department (GF0264).

## Results

### Overall Group

Over the four year period of the UHCW GCA FTP, we found 620 eligible patients. Patients that were less than 50 years old were excluded from this analysis. Patients were predominantly female (411/620; 66%) and White (84%), though 75 (12%) patients are of South Asian descent. The median age was 70 years old (range 50-100, mean also 70.2 years); details are in Table 1. In the whole cohort, 169 patients (27%) received CS for suspected GCA whilst awaiting assessment, 451 patients did not. Some of these were subsequently diagnosed as GCA (37 patients), and CS were continued. Headaches were the commonest reason for presentation (n=451, 73%), with visual disturbance including blurring of vision being the second commonest (n=282, 45%).

**Table 1:** Demographic and ethnicity details for patients on the GCA pathway and clinical diagnosis of GCA.

|  | Year | 2014 | 2015 | 2016 | 2017 | Total |
| --- | --- | --- | --- | --- | --- | --- |
| **Total cohort** | No. patients | 90 | 147 | 119 | 264 | 620 |
| Age | Median age<br>(years) | 73 | 70 | 71 | 70 | 70 |
| Gender | Female | 65 (72%) | 99 (67%) | 81 (68%) | 166 (63%) | 411 (66%) |
| Ethnicity | Caucasian | 80 | 115 | 107 | 219 | 521 (84%) |
|  | S Asian | 7 | 19 | 10 | 39 | 75 (12%) |
|  | Black | 1 | 4 | 1 | 3 | 9 (1%) |
|  | Other | 2 | 9 | 1 | 3 | 15 (2%) |
| **GCA patients** | Numbers | 35 | 30 | 25 | 53 | 143 (23%) |
| Age | Median age<br>(years) | 76 | 76 | 75 | 74 | 75 |
| Gender | Female | 26 (74%) | 20 (67%) | 15 (60%) | 31 (58%) | 92 (64%) |
| Ethnicity | Caucasian | 32 | 26 | 23 | 48 | 129 (90%) |
|  | S Asian | 2 | 2 | 2 | 4 | 10 (7%) |
|  | Black | 0 | 0 | 0 | 0 |  |
|  | Other | 1 | 2 | 0 | 1 | 4 (2%) |
|  | Permanent visual loss | 7 (20%) | 3 (10%) | 6 (24%) | 13 (25%) | 29 (20%) |

### Patients with GCA

Final diagnosis of GCA was made in 143 (22%) patients within the whole cohort. Of these, 92 (64%) patients are female and 129 (90%) are White. The median age was 75 years old (range 52-95). Their demographic details are demonstrated in Table 1.

Very few patients from Black, Asian and minority ethnic groups (BAME) were given this diagnosis (14 patients overall), US was positive in 5 of these, and biopsies were negative in the 3 patients where this was performed.

### Unaffected patients

We were able to discharge 477 (77%) patients referred to the FTP as they did not have GCA. In view of the retrospective nature of data collection, the notes were also reviewed to see if they were subsequently diagnosed to have GCA as a number of patients were discharged at first visit – none of these patients were subsequently given a diagnosis of GCA. Complete data regarding CS use was only available for 112 of the 132 unaffected patients who were exposed to CS. The median duration of CS exposure before US was 2 days and often these patients had come through other secondary care departments or had been admitted to hospital under medical team and then referred to rheumatology which created delays. Overall CS were given for a median of 30 days (range 1-412 days, standard deviation 78 days) – some patients were given a diagnosis of Polymylagia rheumatica, hence the prolonged courses of steroids in some patients.

### Ultrasound Doppler

All patients who attended the UHCW GCA FTP underwent US, those with typical GCA features were labelled ‘positive’. The majority (66%) of patients received their US on the day of their referral. US was performed in 90% of patients within 3 days, and 96% were performed within 1 week – the main reasons for delay were 1) the request was not marked as urgent and not followed up with a phone call to the vascular laboratory or 2) they were requested under the wrong department. The lead vascular scientist audited the images from all vascular scientists and found no significant differences or discrepancies.

Sensitivity of US overall was 50.0% (95% CI 42.9%, 59.1%) (Table 2) compared to clinical diagnosis and improved further in patients with no CS exposure to 55.8% (95% CI 44.7%, 66.4%. In patients who had been started on CS prior to US, sensitivity was 45.5% (95% CI 34.0%, 57.4%). There was excellent correlation between positive ultrasound and clinical diagnosis of GCA. Patients already on CS still showed halos in 12-15% within the first 6 days, and no definite halos were found from day 7 onwards, although only 4% of patients were in this category. Specificity was less affected by CS exposure and the figures were well above 90% in all the groups.

**Table 2:** Analysis of performance of US comparing with corticosteroid use. Performance of US compared to clinical diagnosis.

|  | *All patients*<br>*(612)* | *No CS(443)* | *CS started pre*<br>*US (169)* |  |
| --- | --- | --- | --- | --- |
| *Final diagnosis of*<br>*GCA* |  |  |  |  |
| True +ve | 73 | 43 | 30 |  |
| False -ve | 70 | 34 | 36 | p <0.0001* |
| *Not GCA* |  |  |  |  |
| False +ve | 13 | 12 | 1 |  |
| True -ve | 456 | 354 | 102 |  |
| Sensitivity | 50.0% (95% CI<br>42.9%, 59.1%) | 55.8% (95% CI<br>44.7%, 66.4%) | 45.5% (95% CI<br>34.0%, 57.4%) |  |
| Specificity | 97.2% (95% CI<br>95.3%, 98.4%) | 96.7% (95% CI<br>94.4%, 98.1%) | 99.0% (95% CI<br>94.7%, 99.8%) |  |
| Positive predictive<br>value | 84.8% (95% CI<br>75.8%, 90.9%) | 78.2'' (95% CI<br>65.6%, 87.1%) | 96.8% (95% CI<br>83.8%, 99.4%) |  |
| Negative predictive<br>value | 86.7% (95% CI<br>83.5%, 89.3%) | 91.2% (95% CI<br>88.0%. 93.7%) | 73.9% (95%<br>66.0%, 80.5%) |  |

\* P value calculated using Fisher's exact test, 2 tailed.

Positive Predictive value (PPV) was lower for the year 2015 (74.07%), otherwise the numbers have been consistently above 85%. Overall PPV for US over the 4 years was 84.88%. Negative predictive value (NPV) also seems reasonable at 86.69% which improves further to 91.24% in patients that are CS naïve. As one would expect, negative predictive value was lower for patients already on CS prior to US. False negative scans were seen in 34/443 in CS naïve group and 36/169 in patients already started on CS prior to US - these differences were highly statistically significant (p<0.0001, Fisher’s exact test, 2 tailed). Some US results were indeterminate, and these patients were investigated further usually with TAB if there was ongoing clinical suspicion of GCA. Visual symptoms including flashing lights, temporary or permanent reduction in visual acuity (such as amaurosis fugax), diplopia or field defect had been present in 66 patients at presentation, with complete recovery in 38 patients (58%).

Table 2.

Figure 2.

**Figure 2:**
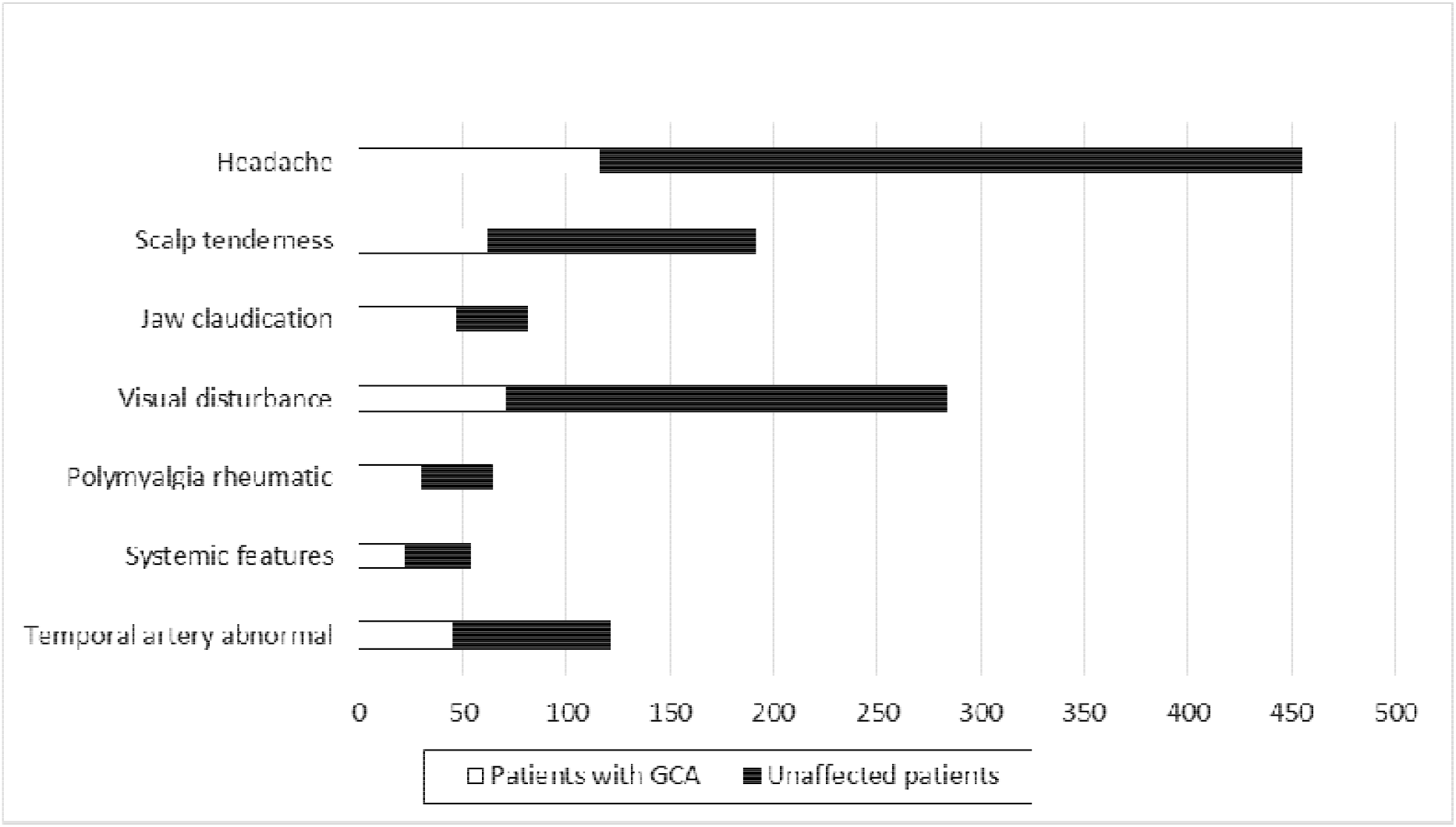
Diagnosis of GCA according to presenting features

### Running the FTP

As the FTP has become more established it has become busier. The number of referrals to the FTP has risen each year though outcomes have remained consistent (Table 2). Although referral rates tend to fluctuate, 2017 was far busier than any of the previous years and this trend is likely to continue. There has been more than doubling of referrals in the 4 years this pathway has been running. This has led to service constraints but we have managed this through better middle grade (core medical trainees, specialist trainees, fellows) support. An internal quality assurance led by the lead vascular scientist did not find any discrepancies and there have been regular multi-disciplinary team meetings to discuss difficult cases and issues.

## Discussion

The world is currently in the grip of a pandemic due to Covid 19 which started in China but has been particularly severe in Europe and US with infections exceeding 35million and deaths exceeding 1 million [14]. Those particularly at risk include ‘people on immunosuppressive therapies sufficient to significantly increase risk of infection’ as outlined by the government of United Kingdom [15]. The global Covid 19 Rheumatology alliance provides some evidence for this assumption with data suggesting poorer prognosis for patients on CS [16].

Corticosteroids are associated with significant adverse effects with some - like psychological side-effects seen within a few days of initiation [17]. Other common side-effects that have led to the development of biological agents such as Tocilizumab [18] include osteoporosis, hyperglycemia and diabetes mellitus, hypertension and infection [19]. CS induced side-effects are quite common with about 80% developing at least one side-effect. Cost evaluation of 7 most common side-effects with CS (fracture, cataract, diabetes, peptic ulcer, stroke, myocardial infarction, non-Hodgkins lymphoma) resulted in extra costs of £165 per person year using CS [20]. Other studies from US have found similar problems with CS [21]. More recent data continue to support the problems with CS use [22]. Hence, there are significant benefits to reducing CS burden both for patients with GCA and for those who do not have GCA. The Coventry FTGCA pathway successfully demonstrates the feasibility of a multi-departmental model of care whereby patients can be assessed and treated urgently, with the additional benefit of minimising use of CS in patients not thought to have GCA. In the current series, >75% of patients who are being referred do not have GCA, so avoiding or reducing CS for these patients is an important achievement.

This pathway is reliant on 3 teams working together to deliver this – vascular physiology scientists, ophthalmology and rheumatology. We have not come across other examples of such team working for delivery of GCA pathways, but there are multiple advantages with this approach.

These include:

1. Vascular scientists are experienced at imaging blood vessels so learning to assess the features of GCA is relatively easy.
2. Vascular scientists can assess vessels other than the temporal artery, such as the carotid arteries and occipital arteries, and produce reports that are acceptable to other departments, for example where there is carotid artery stenosis. Our group has previously published a case series about occipital artery involvement in GCA [23].
3. Patients presenting with visual symptoms including visual loss are assessed by both ophthalmology and rheumatology teams with an integrated management plan with no delay in referrals from one specialty to another. These patients routinely also have carotid US as part of assessment for TIAs/strokes.
4. The pathway is not affected by individual clinician or sonographer absence.
5. Patients can be discharged at the first visit.
6. Reduction in hospital inpatient bed days.

A number of other GCA pathways are run by individual clinicians performing US, which places the system at risk in periods of absence or illness. This pathway also received a commendation from British Society for Rheumatology in 2018 as part of best practice awards. A recent study has shown that even short course of CS have been associated with significant adverse effects [24].

The vascular laboratory is staffed with vascular scientists who are trained to look at blood vessels in different regions of the body. They also provide urgent scans for the Transient ischemic attack (TIA) service. Some patients presenting with visual loss have strokes rather than GCA and these reports with acceptability to other departments are an important element of this service. Despite the limitations associated with multiple team members performing US, we have seen high sensitivity, specificity and predictive values for US within this pathway. These results are comparable to other research studies although often there is a drop off in daily practice compared to results from clinical trials [25,26].

This is the largest reported cohort undergoing US as first line of management for assessment of GCA in the world. Although the median duration of CS exposure before US was only 2 days, there was still a significant drop in sensitivity and US was statistically significantly more likely to be negative for these patients. This has important implications for fast track pathways. We found sensitivity of 55.8% in CS naïve and 50.0% in the whole cohort including patients who had been on CS with specificity of >95% within all groups. TABUL study included 430 patients [27], had sensitivity of 54% and specificity of 81% for US. In the TABUL trial there were 162 patients with US consistent with GCA, 9 of these only had changes in the axillary arteries, inclusion of the axillary arteries increased absolute sensitivity by 5.5%. Some small studies have shown higher sensitivities for US in GCA but with serious methodological limitations [28]. Two separate meta-analyses have been completed to investigate the role of US in diagnosis of GCA [9,29]. For the first meta-analysis, the weighted sensitivity and specificity of the halo sign were 69% (95% CI, 57% to 79%) and 82% (CI, 75% to 87%), respectively compared with biopsy and 55% (CI, 36% to 73%) and 94% (CI, 82% to 98%), respectively compared with ACR criteria. In the second meta-analysis, the pooled sensitivity was 0.68 and pooled specificity was 0.91. Our results are comparable to these, despite the caveats of real life data over a substantial period of time and involving multiple clinicians.

In this cohort, majority of patients with visual symptoms have recovered completely. This is reassuring and supports observations from other fast track pathways that early treatment can result in benefit in patients with visual problems [30]. We had not routinely collected specific data relating to duration of symptoms and delays in referral. Previous studies have highlighted that patients at highest risk of neuro-ophthalmic complications don’t always mount high inflammatory responses [31]. In our cohort, patients presenting atypically to Ophthalmologists with visual symptoms have been included.

### Limitation

This study includes real life data which is not as strictly controlled as clinical trials. Studies of this nature are subject to biases which would be applicable to this as well.

Vascular scientists were only provided informal training; they were not required to provide certification of their skills in this area.

We did not have detail on CS dosing for a number of patients. This may have influenced the utility of US. There may be bias in the way patients were given CS although data do not suggest that.

We only scanned axillary arteries for patients who had arm claudication (very few patients) within this cohort (although we now routinely scan these) – this may have increased US sensitivity by ∼5%. We also routinely performed carotid artery scanning for patients with visual symptoms which is not part of other FTPs.

## Data Availability

We would be happy to provide anonymised data for verification.

## Key messages

1. Coventry FTGCA pathway represents a novel method of delivering care for patients with suspected GCA patients with a collaborative multidisciplinary pathway between Ophthalmology, Rheumatology and Vascular Physiology that has been sustained over many years; this model significantly reduces unnecessary corticosteroid use.
2. Corticosteroid use, even for short duration (median duration was 2 days in this study in patients started on CS prior to assessment) reduces the utility of vascular Doppler.
3. Real life performance of vascular Doppler is comparable to research studies.

## Contributorship

All authors have contributed to the study design, data collection, analysis and write up of the article.

## Acknowledgements

We wish to thank Prof N Krishnan, UHCW NHS Trust and Ines Rombach, Institute for Statistics, University of Oxford for help with statistics and for reviewing the manuscript. We wish to thank Dr K Chaudhuri and all the rheumatology and ophthalmology consultants at UHCW NHS Trust for help with running the pathway, also the vascular surgeons for their help with biopsies and allowing us access to the vascular laboratory.

We wish to thank the Coventry branch of the GCA patient group (PMRGCA UK) for their help and support in creating this model.

We wish to thank British Society for Rheumatology for their support for this model of care.

## Funding info

No specific funding was received from any bodies in the public, commercial or not-for-profit sectors to carry out the work described in this article.

## Ethical approval information

Ethical approval obtained from University Hospital Coventry and Warwickshire NHS Trust Research and Development department – approval number GF0264.

## Data sharing statement

We would be happy to share anonymised data.

## Data availability

Anonymised data can be made available for verification

## Patient and public involvement

The Coventry branch of national GCA patient group (PMRGCA UK) has been involved with the design of this model of this care, and has been very supportive. We did collect some pilot patient feedback, which was very positive, although that has not been reported here.

